# The HR concerns and coping mechanisms for enhancing workplace flexibility in the healthcare sector during the COVID-19 Pandemic: A systematic review

**DOI:** 10.1101/2022.02.03.22270380

**Authors:** Ajit Kerketta, B Sathiayseelan

## Abstract

The COVID-19 pandemic devastated the globe, claimed billions of lives, and hampered trade and regular activities. Safety against the COVID-19 pandemic was the priority of the whole human species in society, the workplace, the family, and personal life. All sectors have been working responsibly to adjust to the new normal. The government is doing its best to implement the best laws and regulations to support and protect livelihood and industry. Healthcare professionals were at risk, and HR employees found it difficult to ensure effective and efficient business operations. The purpose of the study was to examine the role of HR professionals and investigate problems encountered in confronting the COVID-19 pandemic to improve workplace flexibility in the healthcare sector.

The study used an evidence-based technique reproducible from previous research on HR issues in the health industry during the COVID-19 era. Such methodology assisted in producing a wide range of results from within and outside India. The population was selected with the health-related articles published between April 2020 to November 2021 during the COVID-19 era.

The findings showed that redesigning the systems posed the biggest challenge for HR employees and required significant financial commitment, skilled human resources, cutting-edge technology, and an adequate supply of PPE. Thus, the HR managers were tasked with safeguarding a favourable environment by prioritising adequate, efficient, and workplace flexibility.

## Introduction

The COVID-19 pandemic worldwide has caused a significant loss of human life, businesses and regular activities, posing unexpected challenges to sustainable progress ^(1).^ The pandemic halted trade and the hustle and bustle activities of lives for over two years. Every business sector, including the healthcare industry, was working to mitigate the impact of the COVID-19 pandemic but failed to devise practical solutions. It was a struggle for the entire world to create a robust environment at work. Governments, healthcare organisations, and individuals have contributed to a community-wide mitigation strategy to stop the virus’s spread and lessen its fatality rates that hampered social and economic life ^(2).^ Executives in human resources (HR) have also played a key role in developing the global response to build a stable workplace during the COVID-19 pandemic ^(3).^ Resilience is an innate trait considered by individuals’ physical and psychological characteristics ^(4).^ The American Psychological Association (2014) defined it “as the process of adapting well in the face of adversity, trauma, tragedy, threats, or even significant source of stress (para 4) ^(5)^.” The most salient features of HRM challenges stemming from the COVID-19 pandemic involve adjusting new and current health workers to drastically altered work conditions ^(6).^ During a pandemic, clinicians across the globe risked their lives to safeguard others. The world is grateful to all the COVID-19 warriors, especially the medical professionals—doctors, nurses, and other clinicians—for their crucial contribution to healthcare management. Several studies have been done showcasing their work ^(7)^.

However, studies examining the roles of non-clinicians in general and HR staff, in particular, have not been sufficiently studied. For this reason, the current paper attempts to investigate the roles of HR professionals who toiled valiantly during the COVID-19 pandemic to maintain efficient health services. The hospitals and small clinics were inundated with patients. The shortage of human capital, trained health workers, huge financial involvement, obsolete infrastructure, and lack of other resources hampered effective and efficient healthcare operations. The COVID-19 pandemic has rendered the current healthcare infrastructure outdated and inefficient. HR managers tasked with operating the business were naïve about the whole thing. Such a scenario has led to the emergence of resilience as a new tactic ^(8)^, and the healthcare sectors have been entrusted explicitly with adapting to the impending catastrophe. In addition to having access to new technology that stressed them out and posed challenges to the status quo, the HR professionals were also in charge of engaging and retaining skilled and adequate health facilities. They so significantly contributed to placing skilled and qualified health workforce. Therefore, the study aims to investigate the role of HR professionals and the difficulties encountered when implementing actions to curb the COVID-19 pandemic. The study’s findings would deepen the researcher’s understanding of aspirant HR professionals and provide a crucial foundation for further action to address such issues. The research was started in November 2021; however, it underwent several alterations and adjustments that qualified it for publishing.

### Literature review

The COVID-19 outbreak posed unprecedented threats and public health emergency. Healthcare practitioners were under intense pressure to deliver thorough and efficient care and expressed concerns about the healthcare system as well as difficulties for HR professionals. Employees in HR and healthcare were under stress due to the subpar HR strategy, and the healthcare system could not respond adequately ^(9).^ It was challenging for HR managers to maintain existing strategies despite everyday problems, and remote healthcare delivery was not an option. It was challenging to motivate and engage healthcare professionals whose mental health and well-being had been affected and at an increased risk of stress, burnout, moral harm, depression, trauma, and other mental health issues ^(10)^. The composite database incorporated numerous national and sub-national reports to provide the most up-to-date information for a country like India. Discrepancies such as date range overlap or disputes between the federal and sub-national levels resulted in inconsistent and unstable recommendations, posing another problem for administrations ^(11).^ According to World Health Organisation (WHO) guidelines on “Workplace Risk Assessment,” the COVID-19 pandemic is mostly spread through respiratory droplets and contact with contaminated surfaces. As a result, occupational exposure could occur at any workplace. During work-related travel to and from a location with local community transmission.

Contaminated objects or surfaces constantly expose a risk. COVID-19 virus could be transmitted to anyone who comes into close contact with someone infected. Therefore, the probability of COVID-19 virus transmission through human contact is higher ^(12).^ The danger of COVID-19 infection among medical professionals requires effective administrative and HR strategies to address and thoroughly investigate the challenges. Another WHO recommendation combines human resources for health managers and policymakers at the national, sub-national, and facility levels to develop, manage, and sustain the health workforce required to combat the covid-19 pandemic, maintain vital health services, and safeguard the public. HR practitioners confronted substantial difficulties in putting existing ideas into practice. During the covid-19 epidemic, high turnover rates, staff shortages, record compliances, and health workers’ safety, training, and development were all issues in the healthcare industry ^(13).^ Above all, HR staff had to deal with workplace stress, a problem at work, and a threat to physicians’ lives, all while alleviating COVID-19 dread at work ^(14).^ Frontline healthcare workers who were victims of violence, mental stress, and verbal and nonverbal threats faced similar issues. People were frightened of coming into close contact with one another and behaved like “newer untouchables”^(15).^ The isolation of travellers, non-travellers, and infected and non-infected individuals slowed the COVID-19 infection’s spread to some extent ^(16).^ It was a most challenging period for HR staff. The world was confined in itself, and personnel associated with healthcare-related professions were at the forefront, risking and endangering their lives to ensure the lives of others. The pandemic impacted people, infrastructure, finances, shortages of personal protective equipment, hospital upkeep and growth, supplies and logistic support, types of equipment, and many other healthcare devices ^(17).^ Undoubtedly, healthcare personnel faced psychological, physical, mental, and emotional issues provided additional difficulties for HR managers, who needed prompt action.

### Research objectives

1. To investigate the functions of HR professionals performed during the COVID-19 pandemic to ensure workplace resilience in healthcare facilities?
2. To identify problems and quick-fix solutions designed to mitigate the effect of the COVID-19 epidemic.

## Materials and Method

The reviewer searched online for the appropriate methodology used in published systematic reviews (SR) and meta-analyses (MA) that were relevant to the medical field. Additionally, we followed guidelines that apply to all SR phases in research. However, we integrated all these techniques to conclude and create a thorough flow diagram outlining the SR steps. The researcher, however, adhered to the 2020 updated version of the widely used PRISMA guidelines to minimise potential biases ^(18).^ The systematic review is a widely used methodology established in the 1990s to support well-informed decision-making for the health sector, initially focusing on synthesising quantitative evidence from randomised control trials ^(19, 20).^ Since then, systematic review studies have become a common practice in all disciplines ^(21)^. Now, therefore, is a standard practice to incorporate observational studies and qualitative research in systematic reviews ^(22)^. Cook et al. also addressed this relevancy issue by creating the “SPIDER” search engine, specifically designed to locate qualitative and mixed-method studies ^(23)^. Therefore, the findings of the current study are consequently presented in a themed fashion and are more qualitative in nature. This framework is supported by the Cochrane Collaboration and identified components of non-clinical evidence in evidence-based medicine ^(24)^. The review protocol for the current study is still in place and has not been registered/published.

### Search strategy

The search procedure is an important phase in a systematic review study that enables fetching a collection of information from which conclusions may be drawn with the least degree of bias. However, the current study split its search approach into two phases: The first phase of the data search was completed using the manual-searching method ^(25)^ during the designated period of November–December 2021. A detailed and precise search approach integrating various databases is necessary to find relevant research and reduce the possibility of publishing and language bias ^(26)^; as part of several searches, a systematic review of reference lists of other studies or conduct manual searches to identify additional primary studies. Although these procedures are very different and produce specific results, checking the reference list and executing hand searches are frequently regarded as equivalent ^(25)^. The reviewer used the Google scholar database with keywords like “HR problems during COVID-19,” “workplace resiliency,” and “COVID-19 posed obstacles in healthcare” to get the most accurate and pertinent data. The author used three techniques to reduce the bias of manual searches: seeking references from included studies/reviews, extracting author-based articles, and looking for similar publications/cited papers from the indexed lists ^(27)^. The study’s second phase was revised in January and February of 2022 using the Rayyan database and transferred via RIS to notepad under the heading “COVID-19: The HR challenge.”

### Inclusion criteria

The eligibility criteria are based on the topic, study design and date. Articles were published during the COVID-19 era emphasising the function HR personnel played in fostering inclusive workplace resilience. The works of literature must have been published in English between April 2020 until November 2021. Literature that dealt with the topic but did not relate to the COVID-19 era or the healthcare industry was eliminated.

### Study selection

The systematic review’s findings are shown in Fig. 1. The cumulative literature generated 1,118 results, of which 481 were duplicates, 419 were recorded ineligible, and 24 were articles that were eliminated for various reasons. In accordance with the PRISM standards, the primary reviewer self-reviewed the papers to determine their eligibility at the title, abstract, and full-text levels to avoid veering off topic and coherently summarise the result. The research that matched the topic and goals were chosen for further research ^(28)^. Following the elimination of ineligible studies, 257 were the title and abstracts screened. The subordinate reviewer re-reviewed the excluded study to avoid missing the relevant and appropriate paper. After reaching an understanding with the secondary reviewer over an article’s acceptability, the study’s following steps were carried out for full-text analysis. The procedure was repeated for 81 articles with a full-text screening, and the results were accurate and applicable to the goals.

**Figure 1:**
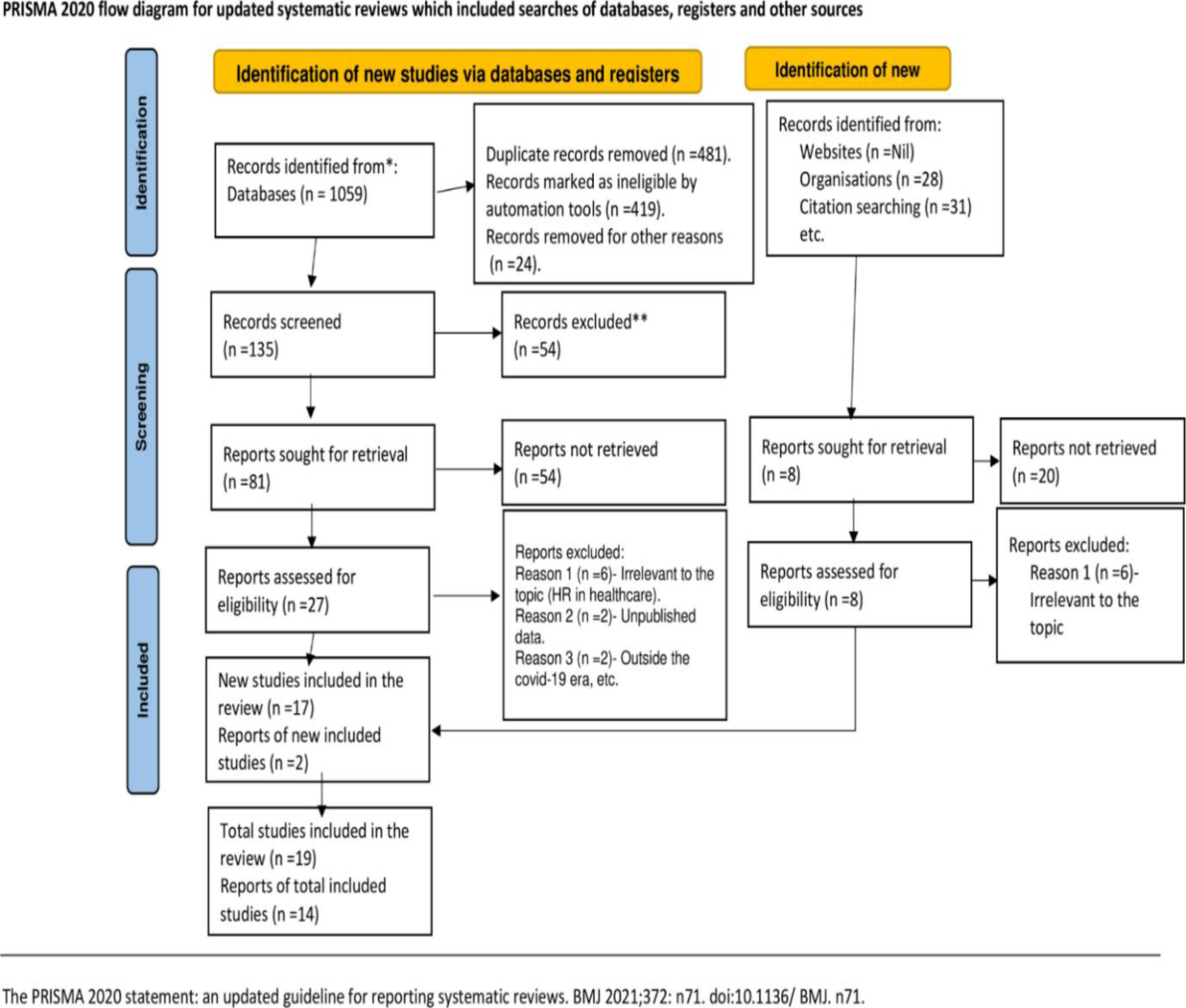
PRISMA 2020 diagram.

### Data extraction

The data were extracted independently. The appropriate evidence was exported from the Rayyan RIS to notepad, and publications that did not fit the inclusion requirements were disqualified from further consideration. Two independent reviewers screened the title and abstract and analysed the content. The PRISMA flow chart (figure 1) is used to report on the entire literature selection procedure. The PRISMA checklist was used to guide the report writing of the systematic review ^(19)^. After full-text data extraction forms, the reported causes posing challenges for HR professionals during the COVID-19 pandemic and their solutions to strengthen workplace resilience in healthcare facilities were gathered.

### Risk of bias assessment

The primary researcher alone conducted the study’s screening, which increased the likelihood of biases. Reviewers met and rigorously and closely investigated the entire work to assess the literature’s quality and minimise the maximum business before coming to a consensus. Furthermore, adhering to the three fundamental principles of study design—participant compatibility, participant selection, and exposure assessment—helped significantly reduce the probability of bias ^(29)^.

### Data synthesis

From the pertinent full-text review, the search approach turned up 27 relevant papers. Since there are not enough publications to categorise with similar characteristics, the data were combined into one group and synthesised individually. However, the reviewers examined each article extensively, keeping the topic in mind, focusing on the health sector, HR challenges, new solutions, and workplace resiliency ^(30)^ in line with the review’s objectives. The findings are summarised in Table 1 and are split into the following four categories: The difficulties faced by HR professionals in column 1, followed by how these difficulties manifested in column 2, the strategies implemented displayed in column 3, and column 4 demonstrate the three systems-levels which parties to the all.

#### A. The challenges encountered during the COVID-19 pandemic

##### Disease System of Early Alert

An early warning system ^(31, 32)^ is a tool for informing healthcare professionals about impending hazards before they occur. It helps reduce the likelihood of harm and allows enough time to protect oneself and others. The early warning system works like an alarm clock, awakening people or groups of individuals at predetermined times, demonstrating readiness ^(33).^ However, ensuring the safety and security of healthcare workers is critical for an organisation’s efficiency in controlling expenses and improving revenue without jeopardising service quality. Therefore, health professionals should be given job security to attain this goal ^(34)^.

##### Finances

The financial crunch during the COVID-19 ^(35, 36)^ was inevitable. Healthcare organisations strive for quality, accessibility, and a scalable business model under the COVID-19 scenario. Trying to figure out the challenges and strategies for dealing with this complex, highly sensitive, and high-uncertainty situation, especially during a pandemic. However, a recent study by Iranian start-up enterprises looked into the impact of the financial crisis on new businesses and found it to be a significant barrier ^(37)^. Allocation of funds for development and expansion from the healthcare sector’s overall budget and financing for the COVID-19 pandemic response has impacted building an adequate system to maintain and build a viable and resilient workplace for healthcare employees.

##### Employee Handbooks

The updating employee handbook ^(38, 39),^ like many other countries, India lacked formal and comprehensive policy updates for healthcare workers that can encourage, mobilise, and assist them during prolonged physical, mental, and emotional weariness, such as the current COVID-19 pandemic crisis. Until now, national and subnational policy recommendations for preventing transmission and protecting workers from the COVID-19 pandemic have been inconsistent. By maintaining an open and safe workplace, policies focused on protecting workers prevent the virus’s spread and protect the health workforce ^(40)^.

##### Healthcare services management

Healthcare service operation ^(41, 42)^ during the COVID-19 pandemic was crucial. Fear, stigma, misinformation, and mobility restrictions have disrupted the resilient workplace and delivery of healthcare services in all situations, posing severe difficulties to the global health system. As a result, healthcare companies must keep the public’s faith in the system’s ability to offer a safe and necessary employment environment while reducing infection risk in health facilities. World Health Organization’s vibrant advice is to ensure appropriate care-seeking behaviour and adherence to public and healthcare worker safety ^(43)^.

##### Management and the healthcare workforce

The global health sector faces a severe dilemma as the COVID-19 epidemic spreads rapidly among healthcare personnel. Managers ensure that the health workers are safeguarded and extend appropriate working conditions. WHO advised to pay remuneration and incentives promptly and on time ^(43)^.

##### New Technologies

The advanced and new technologies ^(44, 45)^ helped the management of the COVID-19 pandemic. However, the new technologies assisted in making timely, evidence-based choices, that healthcare organisations must implement surveillance systems and health information technology to track specific health workforce metrics ^(43)^.

##### Administration and direction

Most organisations are operating on a digital platform during the COVID-19 pandemic; as a result, there is a lack of continuous business activities, employee safety, and consumer preferences, which HR managers are working hard to normalise ^(46)^. The most complicated and crucial resilience-building component for maintaining a resilient workplace in any health system is administration and leadership, often known as stewardship. According to the World Health Organization, management and leadership are linked to the government’s role in health and its relationships with other players involved in healthcare delivery ^(47)^.

##### Health-related human resources and funding

Human resource management ^(48, 49)^ is one of the most critical factors affecting the healthcare system’s performance. HR’s function in health and health funding is to ensure that local health organisations’ human resources are adequate to satisfy the demands of healthcare reform ^(50)^.

##### Human resources shortage

Maintaining an adequate healthcare workforce ^(51, 52)^ is critical to ensuring a safe working environment for healthcare professionals (HCPs) and safe patient treatment. Due to HCP exposures, illness, or the necessity to care for a family member at home as the COVID-19 pandemic proceeded staff shortages. Staff recruitment, turnover, retention, training, and development were all negatively impacted during the pandemic. Other issues that wreaked havoc on healthcare employees were safety concerns, violence, stigma, and digitalisation ^(53)^.

#### B. The components for a health workforce resilience in low-resource management

**Table no. 1:**
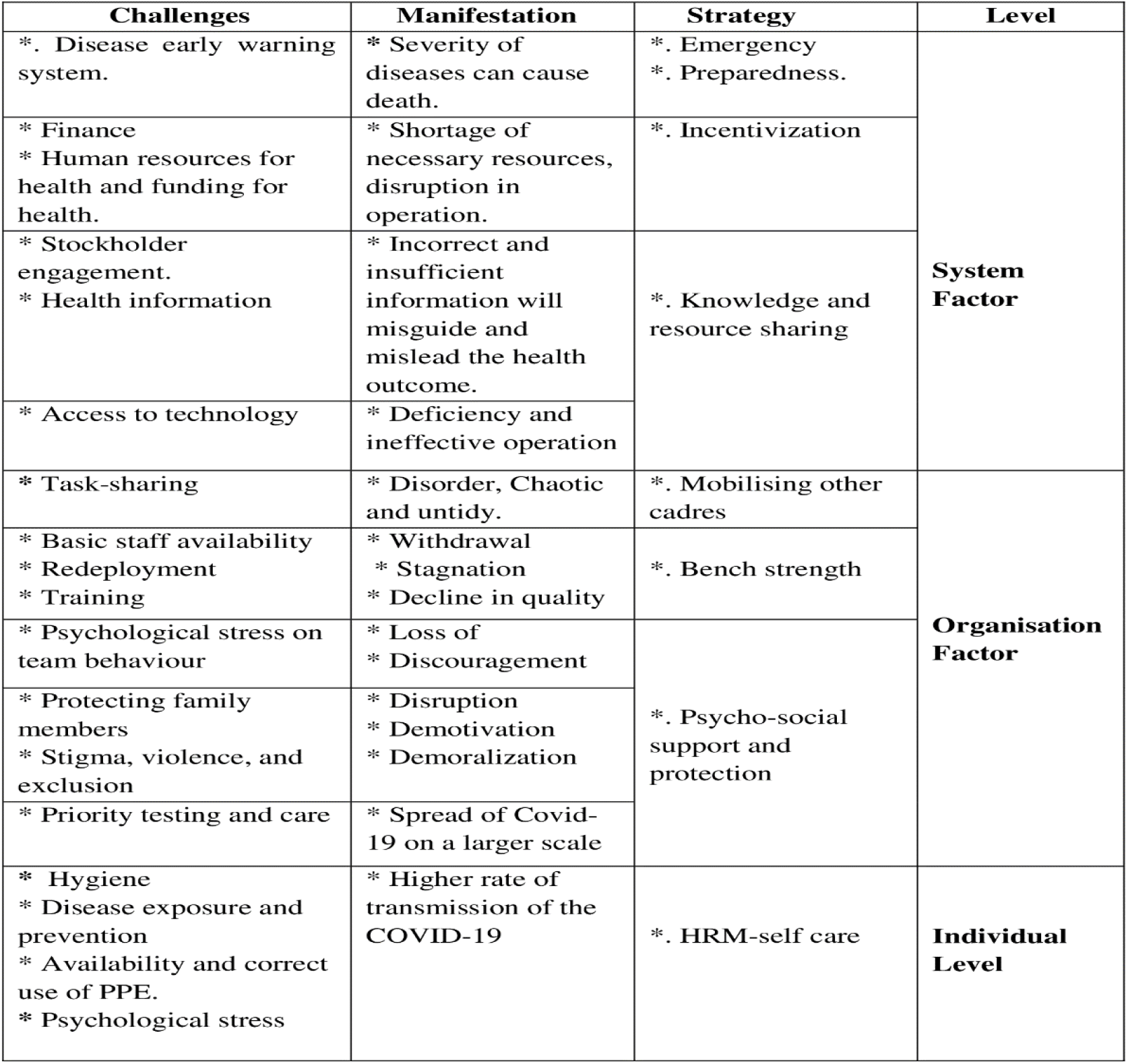
Demonstrates the challenges, their impact, and the coping strategies, categorising them at three levels of influence.

#### A. Levels of variables: At an individual level-workplace resilience

##### Self-Care in Human Resource Management

The key components of infection prevention and control (IPC) measures ^(54, 55, 56)^ suggested personal protective equipment (PPE), as well as suitable equipment and self-isolation and keeping distanced from the infected person.

#### B. The organisational aspect-workplace resilience

##### Support and safety on a psycho-social level

The frontline staff in COVID-19 wards were strengthened by the safety and support provided by psycho-social management ^(57,58,59)^. Enhancing process organisation, enforcing appropriate behaviour, and distributing the responsibility of each all contributed to creating a productive workplace ^(60)^.

##### Enhancing the availability of health workers

Policymakers, executives, administrators, HR managers, and trainers should plan for caseload surges and determine how to mobilise additional health workforce capacity. Determine crucial COVID-19 workforce requirements by quantifying task and time expenditures and combining them with epidemiological Context and demographic data. Identify where to obtain more health workers redeploy existing ones, and establish infrastructure and health personnel. However, providing training assists health workers in developing skills or retooling for the new normal, including exploring realistic options that help build a workable workplace ^(61)^.

##### Organising the healthcare workforce

The health industries must identify the most critical processes within their organisations’ geography and functional structure for further re-evaluation involving management and employees ^(62)^. Because COVID-19 has varying degrees of impact on the global health system that may change over time and temporary staffing may be beneficial, most medical, nursing, and healthcare organisations trained workers and interns to replace the employment deficit, ensuring that vital health services were available and not disrupted. Also, to respond to the COVID-19 pandemic, redeploy employees from non-affected or low-intensity areas inside the health facility or other clinical fields.

### At the system level-workplace resilience

#### Preparedness for a disaster

Imagine something unpleasant and unexpected; the healthcare organisation must prepare and plan for it ^(63, 64)^.

#### Mechanism for Incentivizing

By providing financial incentives, medical professionals and health workers should be recognised for putting their health in danger, being exposed to the COVID-19 pandemic, and caring for transmitted and suspected patients ^(30)^.

#### Governance and cross-cutting cooperation

Hospital cost structure and the implications on cost management during the COVID-19 ^(65, 66)^ mandated that the policy-making process be streamed and national health systems or imposed conditionalities under neoliberal governance must be strictly followed. Many hospitals, notably private hospitals, decided to cut labour expenditures, which caused conflict among the workforce providing patient care. Administrative and executive officials have had to consider alternative measures to keep and sustain health professionals.

## Discussion

Addressing the current circumstance (Covid-19) was one of the most challenging issues for HR professionals ^(67)^ and posed mental, physical, and psychological stress and raised serious concerns about well-being. However, embracing advanced technology solutions and personal protective equipment (PPE) and redesigning the framework provided security and protection to health workers in their workplaces ^(68)^. Nevertheless, based on reviews and first-hand information from frontline healthcare professionals exposed to COVID-19 and other psychological, emotional, physical, and mental challenges, this study offers practical recommendations for building workplace resilience among frontline employees. Within three basic levels, (1). Individual Level, (2). Organisational Level, and (3). System Level, the recommendation comprises a wide range of minor and extensive interventions ^(69)^. The HR challenge and their efforts to create resilient working circumstances, the COVID-19 manifestation, and strategy to control COVID-19 and build a workable working environment revolve around these three variables. To the best of the reviewer’s knowledge, this is the first study of its sort, focusing on HR-related problems during COVID-19, identifying threats, and assessing the measures employed by HR managers to create workplace resilience. A more extensive study demonstrates that all healthcare and non-healthcare workers created a viable work environment. It does not just focus on HR managers’ contributions ^(9)^. World Health Organization has published guidelines, including “Getting your workplace ready for COVID-19” pandemic (69), “COVID-19: Occupational health and safety for health workers” ^(13)^, and “Health workforce policy and management in the context of COVID-19 pandemic response,”^(70)^ “Pandemic resilience and health system preparedness: lessons from COVID-19 for the twenty-first century” ^(71)^ highlighted challenges and strategies the HR personnel adapted to mitigate the spread of the COVID-19 pandemic. The organisational-level responses to the COVID-19 outbreak illustrate challenges, strategies and frameworks in academic institutions adopted to facilitate handling such a crisis shortly at individual and organisational levels ^(72, 73).^

### Limitations

The study’s fundamental flaw is that its ongoing development has disadvantaged researchers. Therefore, it’s possible that the current analysis only found a few studies. However, the researcher attempted integrating as many papers as possible to get the best results. There may be inconsistencies in the study’s conduct that prevent it from producing acceptable and accurate results; however, professional assessment and observation have given the study its expected results. The current study focused on human resources (HR) professionals who were crucial in maintaining a positive work environment in hospitals and ensuring enough resources for the staff members—including physicians, nurses, lab technicians, pharmacists, and operational management—to do their jobs. They were less likely to contract an infection, but managing patients, family members, medical staff, and administration left them mentally and physically worn out. The study has addressed healthcare professionals’ mental, physical, and emotional status, leaving a gap for research on the mental, physiological, and emotional status of non-clinicians, in particular, who experienced the COVID-19 pandemic. As a result, a similar study can be carried out on various non-clinical health professionals including administrators, quality managers, ambulance drivers, and cleaning staff, to ascertain the effects of the pandemic they encountered.

## Conclusion

The Covid-19 pandemic, raging for a long time, has shaken the healthcare system. Ebola, SARS, MERS, and COVID-19 had already confronted the world. Despite this, the healthcare system has been unable to contain the COVID-19 pandemic. The HR initiatives, management policies, and resource availability have failed at every level. The health workforce was subjected to outcries as ordinary citizens and health providers. The COVID-19 outburst was so brutal that healthcare workers had to choose between saving themselves, their families, close relatives, or other patients. During the harsh COVID-19 working conditions, healthcare workers should be supported in a different method to prevent and mitigate any harmful repercussions. To rejuvenate and refresh from stress and exhaustion, the HR manager had to plan out meditation yoga sessions and provide; physical and mental backup that would continue for more extended periods on the job.

## Data Availability

All data produced in the present work are contained in the manuscript.

## Author Contributions

**Author**^**1**^: Conceived and designed the review, data extraction, literature review, performed analysis, and manuscript.

**Author 2:** literature review, reviewed manuscript.

## Conflict of interest

The author declares no conflict of interest.

## Acknowledgements

The author would like to acknowledge Dr Raghavendra A.N. for assessing the methodological quality and helping extract and review the pertinent literature.

## Notes

### Competing Interest Statement

The authors have declared no competing interest.

### Funding Statement

NO. The funders had no role in study design, data collection and analysis, decision to publish, or preparation of the manuscript.

### Summary of Updates

The title of the manuscript was modified. The manuscript's approach was inconsistent in its earlier versions, while the current version strictly adheres to PRISMA guidelines. The figure 1 was updated to PRISMA 2020 flow diagram. The author affiliation was updated.

